# Convex approaches to isolate the shared and distinct genetic structures of subphenotypes in heterogeneous complex traits

**DOI:** 10.1101/2025.04.15.25325870

**Authors:** Saikat Banerjee, Shane O’Connell, Sarah M.C. Colbert, Niamh Mullins, David A. Knowles

## Abstract

Groups of complex diseases, such as coronary heart diseases, neuropsychiatric disorders, and cancers, often display overlapping clinical symptoms and pharmacological treatments. The shared associations of genetic variants across diseases has the potential to explain their underlying biological processes, but this remains poorly understood. To address this, we model the matrix of summary statistics of trait-associated genetic variants as the sum of a low-rank component – representing shared biological processes – and a sparse component, representing unique processes and arbitrarily corrupted or contaminated components. We introduce `Clorinn`, an open-source Python software that uses convex optimization algorithms to recover these components by minimizing a weighted combination of the nuclear norm and of the L1 norm. Among others, `Clorinn` provides two significant benefits: (a) Convex optimization guarantees reproducibility of the components, and (b) The low-rank “uncorrupted” matrix allows robust singular value decomposition (SVD) and principal component analysis (PCA), which are otherwise highly sensitive to outliers and noise in the input matrix. In extensive simulations, we observe that `Clorinn` outperforms state-of-the-art approaches in capturing the shared latent factors across phenotypes. We apply `Clorinn` to estimate 200 latent factors from GWAS summary data of 2,110 phenotypes measured in European-ancestry Pan-UK BioBank individuals (*N* = 420,531) and 14 psychiatric disorders.

## 1 Introduction

Complex diseases often present as heterogeneous groups, encompassing multiple subtypes with distinct clinical manifestations, prognoses, and treatment responses. Such heterogeneous groups of diseases, such as coronary heart diseases, neuropsychiatric disorders and cancers, represent a broad continuum of severity and range of manifestations of several clinical symptoms. For instance, several clinical symptom-based subtypes have been identified using data-driven methods in major depressive disorder (MDD), one of the most common disorders worldwide. Within MDD, almost 1500 symptom combinations are possible [1]. Despite phenotypic diversity, these subtypes may share overlapping genetic and environmental risk factors. For instance, genetic and neuroimaging studies demonstrate that MDD is a genetically and biologically heterogeneous disorder involving multiple genes and brain circuits [2]. Sex and age contribute to the heterogeneity of MDD and are associated with differing symptomatology, risk factors, and responses to treatment [3–9]. Understanding the genetic predisposition of such heterogeneous disorders is crucial for elucidating the biological mechanisms driving both shared susceptibility and subtype-specific divergence.

Genome-wide association studies (GWAS) have estimated the effects of millions of single-nucleotide polymorphisms (SNPs) on thousands of diverse disorders in an effort to better understand these phenotypes. The degree to which phenotypes overlap genetically can inform on their underlying biology. For example, the magnitude of correlation between pairs of trait test statistics represented as *Z* scores corrected for linkage disequilibrium (LD) can indicate how similar traits are genetically [10]. Modeling the genetic covariance matrix can be used to model distinct underlying sources of genetic variance per trait. Examples include bivariate causal mixture modeling [11] and genomic structural equation modeling (gSEM) [12]. Both of these approaches model distinct genetic variance components which can be functionally informative depending on the traits considered. Disorder-specific variants may represent more unique biological processes compared to pleiotropic SNPs. However, approaches to date have several limitations. Bivariate causal mixture modeling does not return SNP-level parameters, instead estimating the degree of causal variant overlap between pairs of traits. While a trivariate version of the method has been developed [13], causal SNP properties across more than three traits cannot be modeled jointly. Methods such as gSEM require that a factor structure be specified, meaning that exploratory analyses or prior hypotheses are required. Finally, gSEM works via pairwise genetic covariance estimates, the estimation of which can be computationally intensive where traits are numerous.

For biobank scale analyses, several factor analysis methods based on matrix factorization have been proposed for extracting shared latent genetic components [14–17]. These approaches aim to factorize the matrix of *Z* scores of associations of SNPs with multiple traits into latent components / factors and their corresponding loadings; sometimes enforcing desired properties on the factors and their loadings. Tanigawa *et al*. [14] used truncated singular value decomposition (truncated SVD, tSVD), a reduced rank approximation of SVD, to identify the latent factors in a computationally tractable and interpretable fashion. Zhang *et al*. [15] provided a Bayesian framework for the matrix factorization, `FactorGo`, where prior distributions of effect sizes are specified and factors are estimated probabilistically. Another recent approach, `GLEANR`, was developed to estimate regularized latent genetic factors in the presence of sample sharing [16], by enforcing sparsity on the factors and their loadings. However, existing approaches suffer from two main shortcomings: (a) lack of sensitivity with respect to grossly corrupted or outlying observations, which are an inherent feature of genetic association matrices; and (b) nonconvex formulations, which yield different results at each run. This poses an issue from a reproducibility standpoint; results are sensitive to the addition of phenotypes, variants, or simply refitting from a different initialization.

Here, we circumvent these issues and introduce *Convex LOw Rank Inference via Nuclear Norm*, `Clorinn`, which derives a low-rank “outlier-free” matrix *X* from a matrix *Z* comprising *N* traits and *P* variant Z-scores using rank minimization, as opposed to matrix factorization. Using *X*, standard factor analysis methods can be applied to examine trait loadings, important SNPs, and global phenotype structure with greater latent factor resolution than methods applied directly to *Z*. Additionally, the low-rank matrix factorization approach of `Clorinn` is convex, meaning that our method is robust to large amounts of noise or the addition of extra phenotypes/variants. We demonstrate that `Clorinn` is computationally efficient and capable of identifying latent factors with greater accuracy in the presence of varying degrees of noise than other methods in extensive simulations. We apply `Clorinn` to 2, 110 real traits from the UK BioBank and identify several trait-group specific factors, allowing us to validate the results from `Clorinn` with the existing literature. Finally, we apply `Clorinn` to 14 psychiatric traits and characterize shared genetic latent factors to demonstrate our method’s utility in a more focused setting.

## 2 Results

### 2.1 Method overview

We assume that the noisy matrix of observed *Z* scores for *N* traits and *P* SNPs, **Z** ∈ ℝ^*N ×P*^, can be decomposed as a sum of a low-rank matrix **X** ∈ ℝ^*N ×P*^ and some error **M** ∈ ℝ^*N ×P*^,

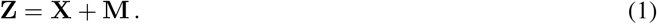

**X** corresponds to the shared genetic components of the traits. The problem to recover **X** from noisy **Z** is called *low-rank matrix approximation* (LRMA). It seeks the best rank-*k* estimate of **X** by solving the problem,

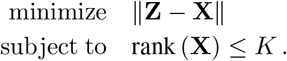

There are two popular approaches for LRMA: *low-rank matrix factorization* (LRMF) and rank minimization.

LRMF seeks to recover latent structures in the data by factorizing the original data matrix **Z** into a linear combination of *K* shared trait-specific loadings **L** ∈ ℝ^*N ×K*^ with latent factors **F** ∈ ℝ^*P ×K*^,

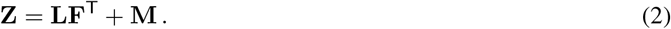

It uses the estimates 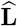 and 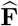 to reconstruct the low rank estimate 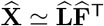. Intuitively, the hidden factors **F** are some unknown underlying latent embeddings of the variants in *K* dimensions. These hidden factors can capture interesting biological insights or pathways that drive complex diseases. The above factorization implicitly assumes that factor-trait and variant-factor relationships are all linear. The number of factors (or dimensions) *K* needed for faithful representation of **Z** will depend on the complexity of the data. As in classical principal component analysis (PCA) [18–20], (2) can be efficiently solved via SVD if **L** and **F** are unregularized and the noise **M** is small and *i*.*i*.*d*. Gaussian. Under this regime SVD enjoys a number of optimality properties. Other common strategies, such as independent components analysis (ICA) and non-negative matrix factorization (NMF) enforce desired properties on **L** and **F**. More recent approaches includes DeGAs [14] and FactorGo [15]. DeGAs applies SVD and truncates the columns of **L** and **F** to obtain low rank. FactorGo uses Bayesian priors on **L** and **F** to enforce sparsity. However, these methods are susceptible to the shortcomings mentioned above. There are two main issues with these existing approaches:

- **Outliers and noise**. One major shortcoming of classical PCA is its brittleness with respect to grossly corrupted or outlying observations [20]: A single grossly corrupted entry in **M** could render the estimated 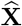 arbitrarily far from the true **X**. These methods assume that the noise is Gaussian, *i*.*e*., **M** *∼* δ (**0, Σ**) is a matrix of Gaussian residuals with variance **Σ**. In many cases, particularly for heterogeneous diseases, the *Z* scores may have non-Gaussian outliers and discrepancies.
- **Non-convex optimization**. With the exception of PCA, matrix factorization models require non-convex optimization, which cannot be fully reproducible. Many practitioners fix the random seeds, which is only a bandaid solution as tiny perturbations in the data may yield very different results.

In this paper, we focus on rank minimization methods to obtain a low-rank, “outlier-free” matrix **X**, before applying the matrix factorization via SVD. In recent computer vision literature, a class of robust PCA methods have been developed to handle outliers and heavy noise [21–25]. Following these methods, we assume that the errors **M** can be arbitrary in magnitude but sparsely supported, affecting only a fraction of the entries of **Z**. The outliers and discrepancies captured by the sparse component **M** may include: (a) the private contribution of a few key variants to specific phenotypes, or (b) measurement or manual errors in the available summary statistics.

Therefore, instead of using Gaussian noise, we assume that **M** is sparse for the rank minimization problem. Since direct rank minimization is itself a nonconvex problem, it is often approximated by minimizing the *nuclear norm* of the estimated matrix **X**, which is a convex relaxation of minimizing the matrix rank [26, 27]. This methodology is called nuclear norm minimization (NNM). The nuclear norm of a matrix **X**, denoted by ∥**X**∥_*_, is the sum of its singular values. NNM approximates **Z** by **X**, while constraining ∥**X**∥_*_. NNM is analogous to Lasso regression which approximates minimizing the number of non-zero regression coefficients.

Here, we use different convex optimization algorithms to perform LRMA:

1. Robust PCA (RPCA) using inexact alternating Lagrangian multipliers (IALM) algorithm,
2. Nuclear Norm Minimization (NNM) using Frank-Wolfe (FW) algorithm, and its variant,
3. Sparse Nuclear Norm Minimization (NNM-Sparse) using FW algorithm.

For Robust PCA, we used Algortihm 5 from Lin *et al*. [28]. We derived novel Frank-Wolfe algorithms for the NNM problem, as described in the Methods section.

Having estimated 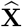 via one of these methods, we use tSVD to obtain 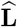 and 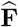 Fig. 1. Our convex algorithms guarantees reproducible estimates of 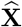. By applying tSVD on 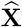 instead of **Z**, we avoid the shortcomings of classical PCA and obtain more robust estimates of the latent factors. In a statistical genetics context, this is especially pertinent; technical noise, measurement error, and population structure can have subtle impacts on association study test statistics. Characterization of latent components representing shared genetic effects via PCA is sensitive to variation arising from these non-biological sources. The estimation of 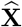 facilitates the description of accurate and reproducible latent genetic factors robust to the presence of outliers, allowing us to mitigate these issues.

**Figure 1:**
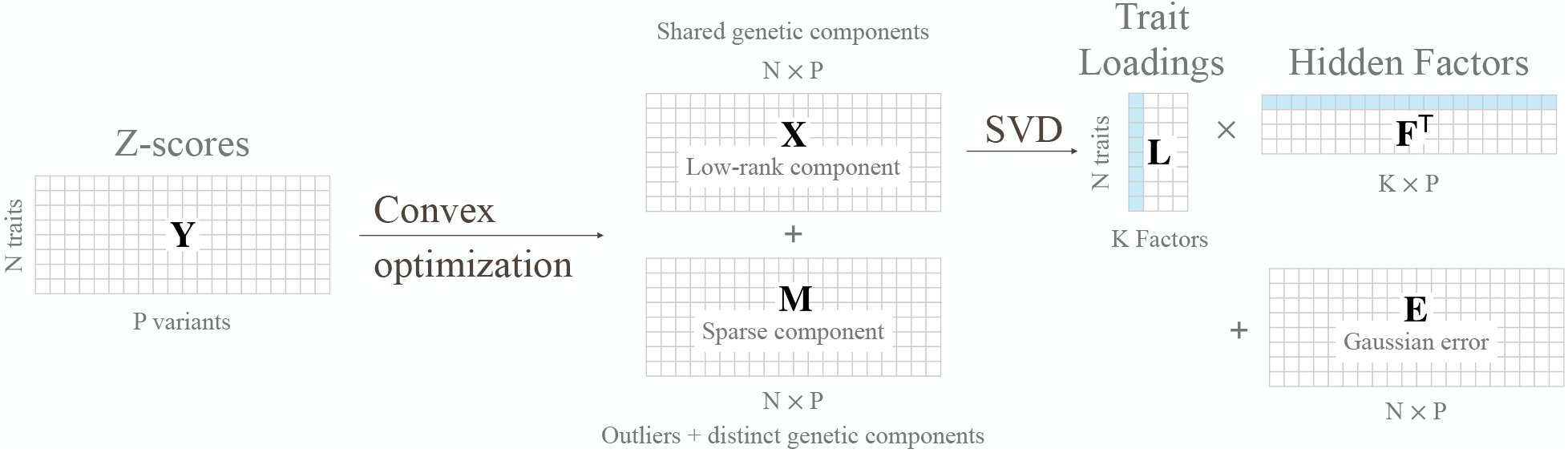
Schematic representation of Clorinn.

### 2.2 Numerical experiments on simulated data

We characterize how well `Clorinn` is able to recover the underlying hidden factors under different conditions and compare its performance to other similar approaches using simulated data. We performed simulations under the model of Eqn 2. We start by generating *K* orthonormal vectors [29] of length *P* (number of variants), which form the columns of **F** ∈ ℝ^*P ×K*^. Each column of **F** (denoted by **F**_:*k*_) is an unit vector with mean 0, standard deviation 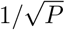 and is orthogonal to all other columns. They represent the underlying orthogonal hidden factors. For each hidden factor, we generate the latent embeddings for *N* studies (traits), which form the columns of **L** ∈ ℝ^*N ×K*^. To mimic the shared component of traits in real data, we sample each column of **L** from a multivariate normal distribution **L**_:*k*_ *∼ N* (**0**_N_, **Σ**) _*n*_; with **Σ** = **(gg**^T^ *⊙* **A)** */K*, where *⊙* denotes element-wise multiplication. Here, **g** ∈ ℝ^*N*^ is a column vector where element *g*_*n*_ is the square root of the heritability 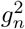 contributed by the low rank component **LF**^T^ to the *n*^th^ trait. The total heritability of the *n*^th^ trait is 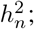 the remaining heritability 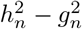 is contributed by the sparse component **M. A** is a block diagonal matrix which encodes the strength of sharing between between different traits within and across disease categories. Each block can be considered as representing a broad disease category with multiple traits. Let *Q* be the number of disease categories (number of blocks) in **A** and *A*_*q*_ be the set of indices for the *q*^th^ block. Then, each element **A**_*ij*_ is given by,

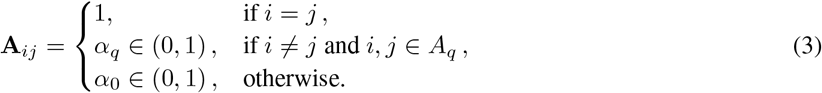

Here, *α*_*q*_ is the amount of sharing between the traits of the *q*^th^ disease category, whereas *α*_0_ is a measure of sharing between the different disease categories.

We then generate **M** ∈ ℝ^*N ×P*^ by sampling each column **M**_:*p*_ from a Laplace distribution,

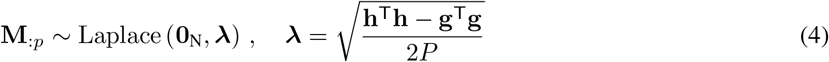

to ensure that the total heritability for the traits is **h**^T^**h**. Each column of **M** is the portion of the *Z* score of a SNP which cannot be explained by shared factors. The true effect sizes are then calculated as,

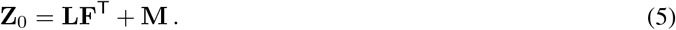

Finally, we add some measurement noise to the true *Z* scores to obtain the observed *Z* scores,

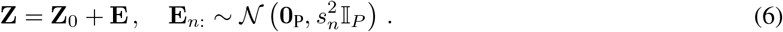

We assume that the variants are independent (i.e., no LD) and that the genotype and phenotype have been standardized, so that the standard error *s*_*n*_ for the effect sizes depend only on the *n*^th^ study (trait). In particular, we can assume 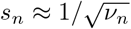 where *ν*_*n*_ is the sample size of study *n*. If the genotypes were not standardized, then the standard error for the effect sizes would also depend on the minor allele frequency of each variant.

An example simulation with *Q* = 3 disease categories is shown in Fig. 2a. The covariance of the loadings is block-diagonal (Fig. 2a right panel) and the loadings of the first two factors show 3 different heterogeneous disease groups (left panel). We compare `Clorinn` versions with tSVD and FactorGo as we vary the number of variants, number of hidden factors and the heritability, respectively (Fig. 2b-d). When the assumed number of factors (10) matches the ground truth, `Clorinn` and tSVD provides better estimates of the factors and their corresponding loadings, whereas FactorGo provides a better estimate of 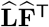. As we increase the number of ground truth factors (keeping the number of inferred factors at 10), all methods get worse at estimating **L** and **F**, but the trend remains similar. The trend however reverses when the number of ground truth factors is less than the assumed *K*. That is, overspecification of number of factors *K* penalizes `Clorinn` more than other methods, as compared to underspecification of number of factors. In the top row, we show the adjusted mutual information score, which measures whether the estimated disease groups match the ground truth. The disease groups were estimated by clustering 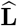. Clearly, `Clorinn` performs much better in recovering the disease groups compared to other methods. This is important as a key goal is to disentangle the different heterogeneous disease groups.

**Figure 2:**
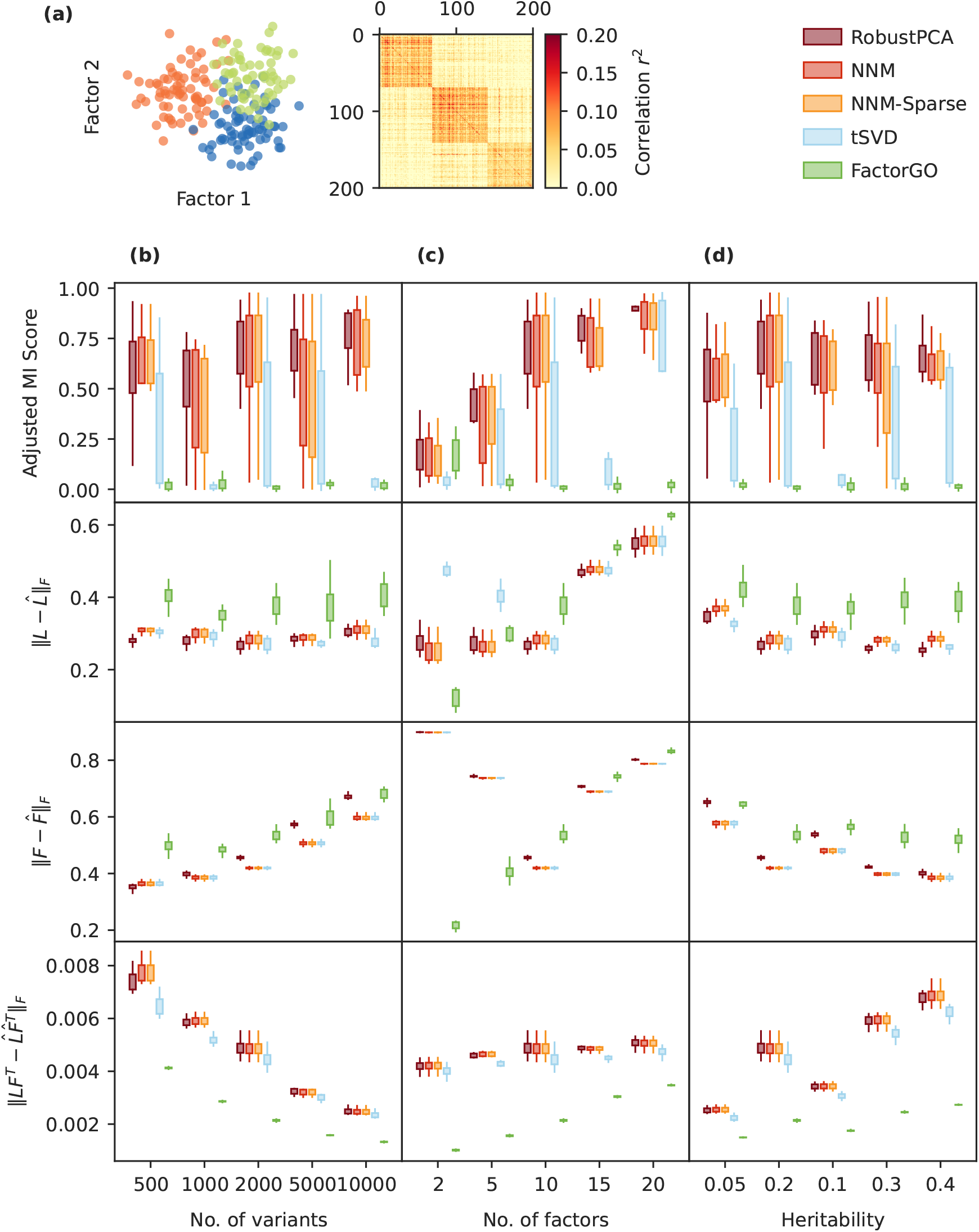
Clorinn accurately classifies subtypes of heterogeneous traits in realistic simulations.(a) Left panel shows the loadings of the first two factors in our ground truth, right panel heatmap shows the covariance of the corresponding loadings. (b) Comparison of the different methods as we vary the number of variants used in the simulations. The comparison metrics for each row are shown on the left. 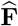 and 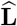 are the estimates of **F** and **L** by each of the methods. Adjusted mutual information score measures whether the estimated disease groups match the ground truth. Each boxplot comprise of 20 simulation replicates. (c) Same as column (b) but with varying number of hidden factors. Each method used *K* = 10 for estimating the hidden factors irrespective of the ground truth. (d) Same as column (b) but with varying heritability of the traits.

### 2.3 Application to 2,110 UKB traits

Having demonstrated the performance in simulations, we used `Clorinn` to analyze 2,110 real traits from the Pan-UKB data. For prefiltering (see Methods) the Pan-UKB data, we followed the same procedure described by Zhang *et al*. [15], but we excluded the prescription traits. We constructed a matrix of GWAS *Z* scores at 51,368 HLA LD-pruned SNP variants across 2,110 traits. We applied `Clorinn` on this input matrix to obtain the low rank matrix 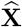. For NNM-Sparse, we used the NN constraint of *r ≤*155872 and *𝓁*_1_ constraint of *l≤* 1770, which were chosen by cross-validation (see Supplementary Figure S1). We then used tSVD to obtain the top 200 factors and their corresponding loadings.

Our first goal was to analyze how the low rank matrix captures the shared genetic components. We observed that the nuclear norm of the low rank matrix, 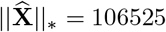, is almost 25% of the input matrix, ∥**Z**∥ = 431234. The matrix rank of 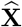 was 560. We classified the 2,110 traits into 30 broad disease categories from their text descriptions, using a publicly available sentence-transformers model (`ls-da3m0ns/bge_large_medical`). We observed that trait clusters obtained from 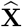 matches closely with their text-based unsupervised classification (Fig. 3). The clustering of traits using tSNE is based solely on their genetic associations, while the color-coded classification in the figure is based solely on their text description. Thus, the genetic association matrix contains similar information about the traits as their text descriptions and allows clustering into meaningful categories. However, we emphasize that the distance between the clusters in Fig. 3 has little meaning.

**Figure 3:**
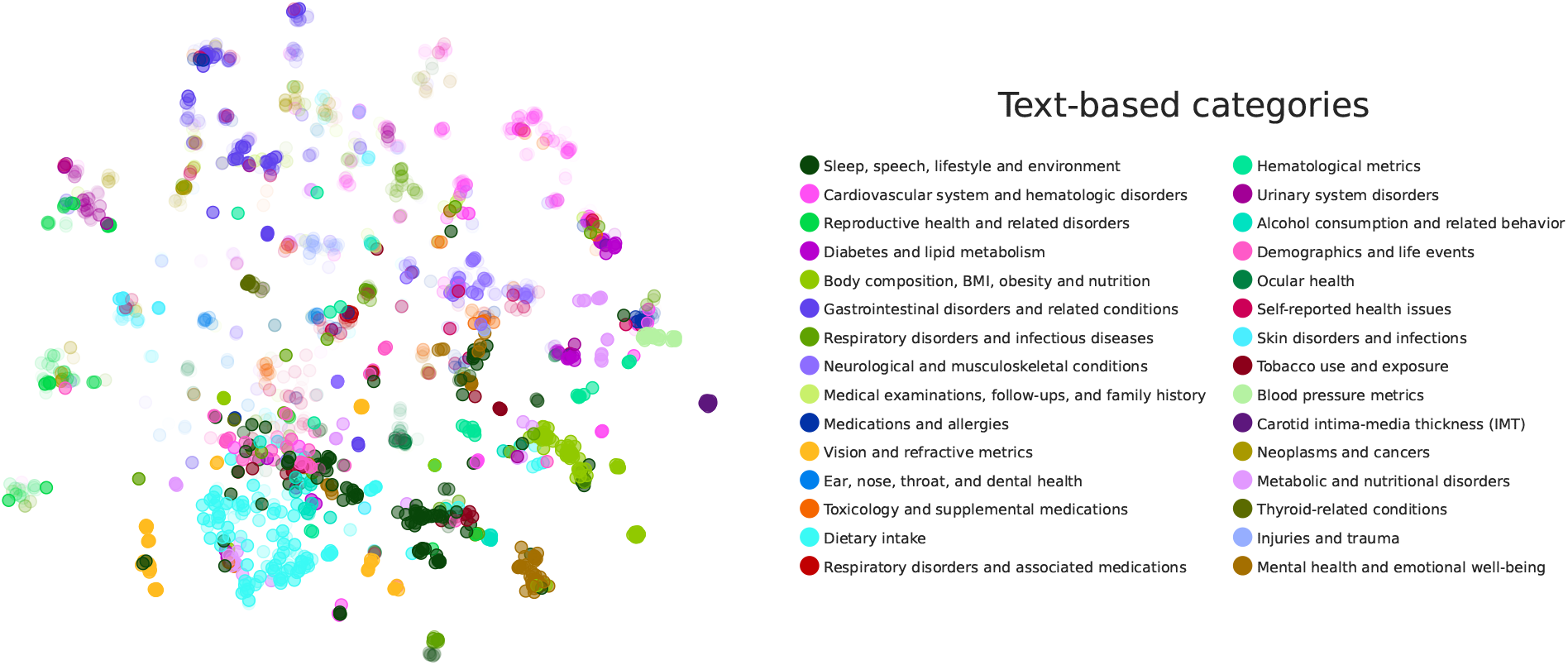
2D tSNE embedding of the Clorinn low rank shared genetic matrix of 2,110 PanUKB traits group into clusters similar to their text descriptions. We classified the 2,110 traits into 30 categories from their the text descriptions, using a publicly available sentence-transformers model. Each point is a trait, colored by its disease category as mentioned in the legend. The opacity of each point is proportional to their estimated heritability.

To illustrate how `Clorinn` captures the shared genetic components of related traits, we characterized the latent components/factors 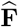 and their corresponding loadings 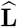; and how they capture related sets of phenotypes, genes and variants in genetic associations. We used the top 200 factors for our analyses. We selected Type 2 diabetes as a focal trait to explore, given its significant genetic heritability. We quantified the relative importance of the factors using squared cosine scores [30]. The top 4 components of Type 2 diabetes (PC2, PC26, PC1 and PC17) explained over 55% of the genetic associations (23.6%, 14.4%, 10.7% and 6.5% respectively). The PCA plots in Fig. 4a and b show the loadings of all traits corresponding to these factors. The top 20 traits for each factor are annotated on the plots (top 40 traits for PC1). We found that height, weight and BMI related traits are the major contributors to PC1 and PC2. PC26 and PC17 are driven by diabetes-related traits and other major risk factors of Type 2 diabetes, including cholesterol levels, indicating that these factors capture the shared genetic components of these phenotypes.

**Figure 4:**
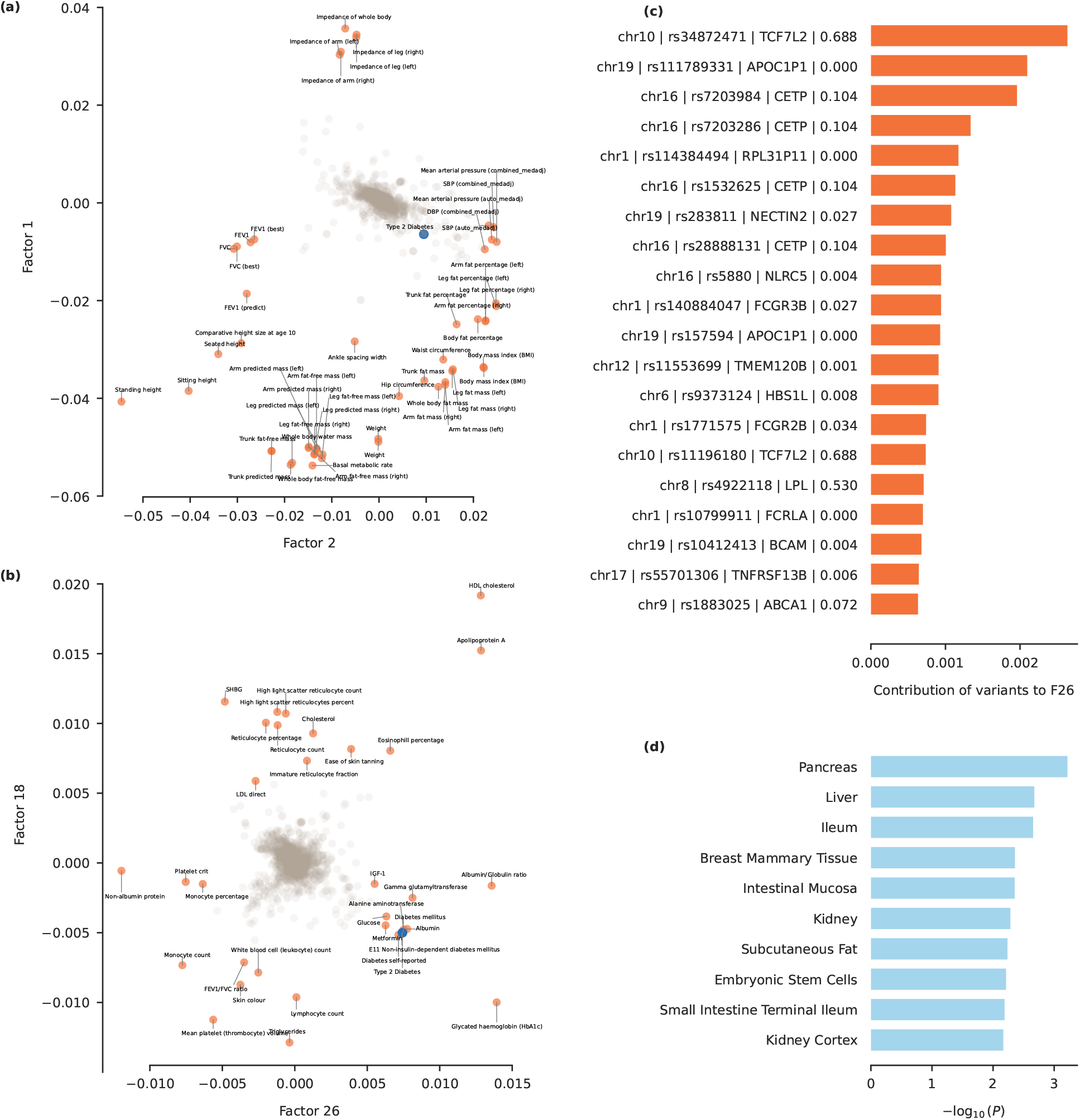
Characterization of hidden factors of Type 2 diabetes, as obtained from the PanUKB analysis. (a) Loadings of all traits corresponding to PC1 and PC2, traits with highest variation are annotated. (b) Same as (a) for PC17 and PC26. (c) Contribution score of variants to PC26. Labels on the y-axis include chromosome number, rsid of the variant, name of the nearest gene and association score of the gene with Type 2 diabetes as obtained from the Open Targets API. (d) Significantly enriched LDSC-SEG annotations for PC26.

Next, we identified the genetic variants contributing to PC26 using the variant contribution score. In Fig. 4, we show the names of theses variants and their nearest genes. Using the OpenTargets API [31], we obtained the association score of each gene with Type 2 diabetes. OpenTargets associations aim to aggregate all evidence referring to the target and disease and the association scores range between 0 and 1, with higher scores indicating stronger associations. We find that many genes that contribute to PC26 (*e*.*g*., TCF7L2, LPL, CETP, NECTIN2, CGR3B) have known associations with Type 2 diabetes, thus establishing consistency with previous molecular findings. Finally, using stratified-LD score regression [10, 32] on 205 tissue– and celltype-specific annotation, we found that PC26 was significantly enriched in pancreas, liver and ileum tissues. This is concordant with the etiology of Type 2 diabetes.

### 2.4 Application to 14 psychiatric traits

`Clorinn` results in a decomposition of the input association matrix into the sum of two matrices, **X** representing ‘uncorrupted’ entries and **M** representing a noise component. This can be conceptualized in a genomics context as a ‘shared’ space and a ‘disorder-specific’ space. We sought to compare application of `Clorinn` to results of a recent analysis of 14 psychiatric traits using gSEM to examine the concordance in latent factor embeddings and solution properties [33]. We obtained GWAS summary statistics for each of the 14 analyzed traits using the best-powered available study (Table 1). SNPs were clumped using a subset of European individuals from the Haplotype Reference

**Table 1:**
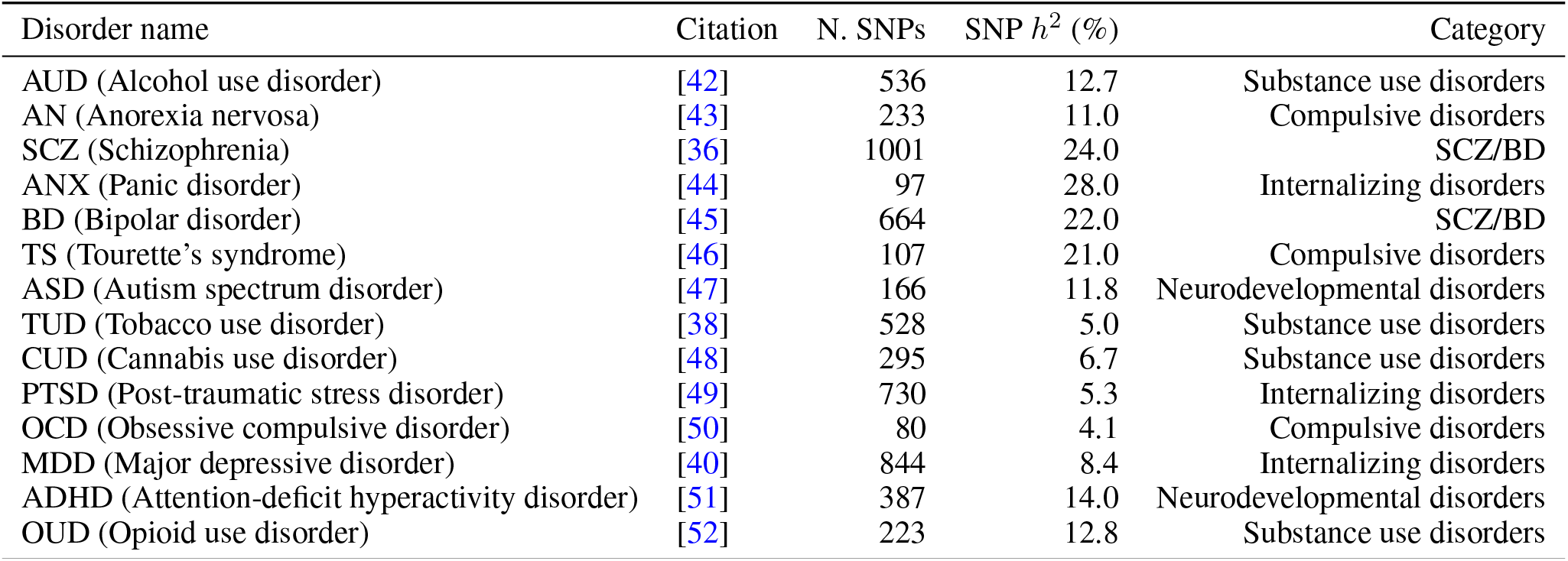
Table of phenotypes included in the cross-disorder phenotype analysis. N. SNPs refers to the number of clumped SNPs in the final pruned matrix contributed by that phenotype. SNP *h*^2^ estimates are taken from the *LDSC* parameter estimate from each respective study in European populations on the liability scale [10]. Phenotype categories are taken from the cross-disorder group analysis [33].

Consortium (HRC) as a reference with a *p*-value threshold of 5*e*^*−*5^. We excluded SNPs with missing association statistics for *≥* 4 traits, resulting in a final matrix **Z** of 14 phenotypes with 5, 815 SNP effects each.

Applying `Clorinn` with *λ* = 0.016 to **Z**, we obtained an **X** matrix with nuclear norm of 867, marking a decrease from the nuclear norm of the input matrix (∥*Z*∥_*_ = 1843). The *λ* value was chosen to yield a **M** matrix with approximately 50% non-zero values. We applied PCA to the resultant **X** matrix and plotted the first two PCs (Figure 5A). We found that phenotypes clustered into the categories assigned from [33]. Further, we observed that the Silhouette score was higher for the first two PCs of **X** compared to that of **Z**, indicating strong clustering via application of `Clorinn` (0.21 vs. 0.06, Supplementary materials).

**Figure 5:**
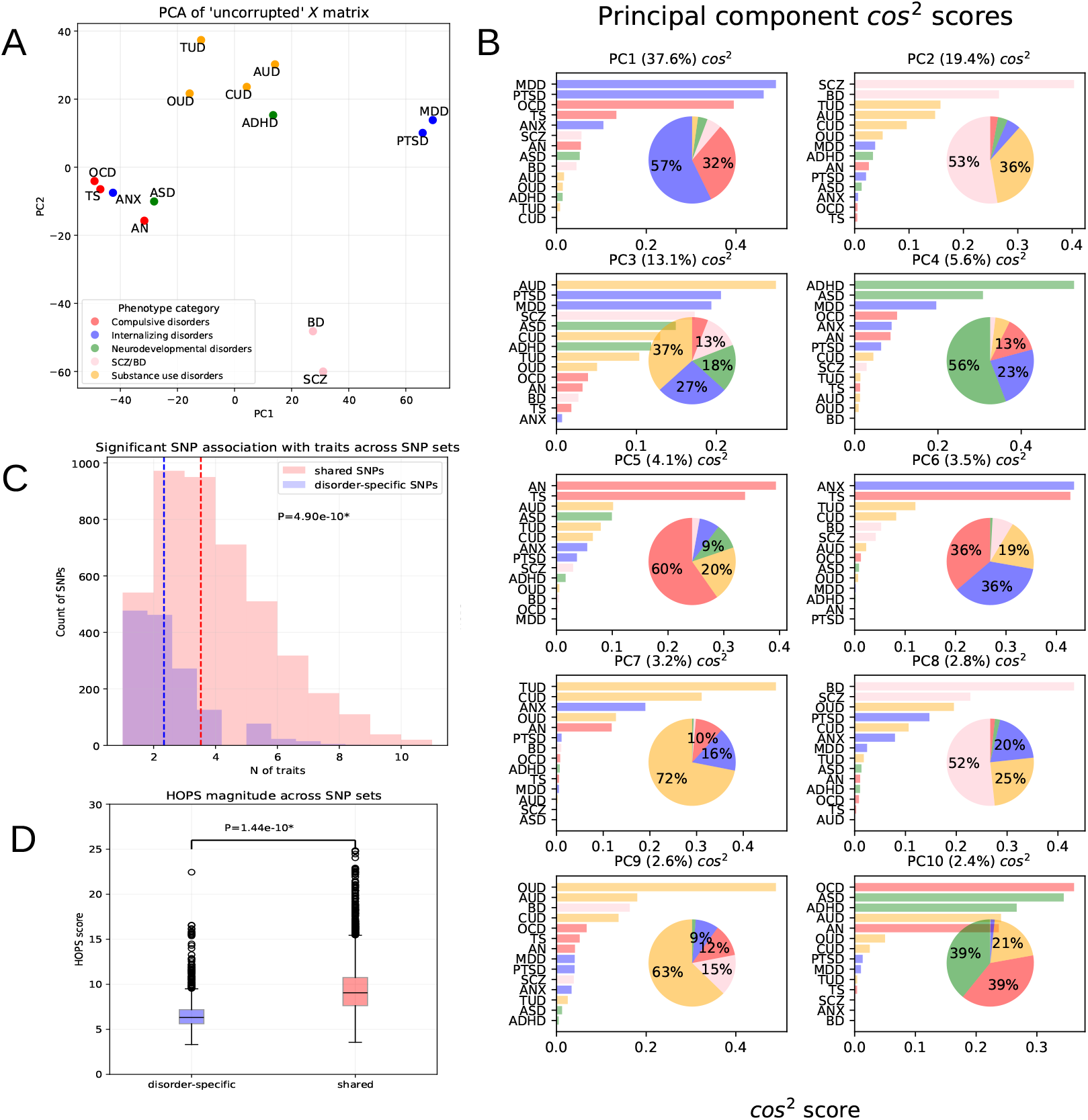
**A**: First two PCs from PCA of ‘uncorrupted’ **X** matrix of 14 psychiatric traits. Phenotype categories are derived from the 5 factor gSEM model from [33]. Phenotypes are labeled by their abbreviations: AUD (alcohol-use disorder), AN (anorexia nervosa), SCZ (schizophrenia), ANX (panic disorder), BD (bipolar disorder), TS (Tourette’s syndrome), ASD (autism spectrum disorder), TUD (tobacco-use disorder), CUD (cannabis-use disorder), PTSD (post-traumatic stress disorder), OCD (obsessive-compulsive disorder), MDD (major-depressive disorder), ADHD (attention-deficit hyperactivity disorder), OUD (opioid-use disorder). **B**: cos^2^ scores per PC with associated aggregated contribution of phenotype categories in pie charts. Category contribution is measured as the normalized cos^2^ score of each phenotype to a PC aggregated by phenotype category, ordered from left to right (ascending). The variance explained by each PC is described in parentheses. The color code corresponds to the legend of A. **C**: Respective distributions of number of traits where SNPs from each set are nominally significant 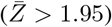. Shared SNPs are those with mean absolute **M** values less than their mean absolute **X** values; disorder-specific SNPs are those with mean absolute **M** values greater than their mean **X** values. Mean values of each distributions are denoted by dashed vertical lines, and the *p*-value was derived from an independent *t*-test of means between distributions. SNP counts are described on the y-axis and number of traits where SNPs are nominally significant are described on the x-axis. **D**: Boxplot of HOPS scores for each set of SNPs, representing their pleiotropy magnitude [34]. The *p*-value was derived from an independent *t*-test of means of pleiotropy magnitude across SNP sets.

The relative contribution of traits to PCs can be assessed using the cos^2^ score. We derived this quantity for the first 10 PCs and aggregated contributions by phenotype category to compare results to latent factor embeddings from previous gSEM results [33]. We find that every factor from the 5-factor model in Figure 2B of [33] is represented by at least one PC in our analysis. gSEM fits a model to a genetic covariance matrix of traits using genome-wide information from approximately 1.6 million HapMap3 variants per phenotype, whereas `Clorinn` approximates the same factor structure in this phenotype panel using 5, 815 SNP effects per phenotype. We visualized the contribution of individual phenotypes to PCs in Figure 5B.

Given the representation of our two decomposed matrices, we hypothesized that differences between the shared **X** and disorder-specific **M** matrices could be informative about SNP properties. We identified 1,539 SNPs where the average absolute value across phenotypes was greater in **M** than in **X**, suggesting these SNPs were more disorder-specific. We expected these SNPs to be nominally significant (*Z* score *≥* 1.95) in fewer traits compared to SNPs with higher values in **X**, which we considered more shared. This was confirmed: disorder-specific SNPs were significant in fewer traits on average than shared SNPs, and this difference was statistically significant (Figure 5C, *p* = 4.9 *×* 10^*−*10^). To more rigorously assess statistical significant, we repeated this operation using 1, 000 random re-samplings of 1, 536 SNPs from **Z** to examine if our observed P-value was due to chance rather than property differences between SNP sets. We found no test statistics more extreme than our observed results in these re-samplings. Similarly, we expected that shared SNPs were likely to be more pleiotropic than disorder-specific SNPs. To examine this, we calculated the HOrizontal Pleiotropy Score (HOPS) [34] in each set across our 14 phenotypes and tested for a difference in means. We found a statistically significant difference in pleiotropy magnitude between SNP sets, with disorder-specific SNPs having a lower mean pleiotropy score than shared SNPs (Figure 5D). Across 1, 000 random samplings of 1, 536 SNPs from **Z** to act as permuted disorder-specific SNPs, we found no test statistic as extreme as the observed *p*-value.

The degree to which PCs load on variables (traits) is given by multiplying eigenvectors by the square root of their eigenvalues. A method to conduct statistical hypothesis testing of loading values was previously introduced in [35]. Briefly, the relationship between raw variable values across samples and the PC value across samples is expected to be *t*-distributed under the null assumption of no correlation; this intuition stems from the fact that larger variable loadings imply stronger correlation of variables to respective PC values. The association of a variable with PC activity can be formally investigated by regressing PC values across samples on input variable values. Following this procedure, we regressed scaled PC1 values on each SNP individually. We found that 18 SNPs had genome-wide significant (GWS) test statistics (*p ≤* 5 *×* 10^*−*8^, Supplementary Note). We expected that these SNPs would be primarily composed of SNPs significant for phenotypes in the internalizing or compulsive disorder categories based on Figure 5B. 14 of the 18 SNPs were GWS for either PTSD or MDD, and 4 were not significant in any of the top 4 phenotypes loaded on by PC1. These SNPs were rs1604060, rs2196150, rs2865303, and rs865020. rs1604060 and rs2196150 were both included as clumped SNPs associated with SCZ [36], with rs2196150 also previously reported as a significant association of leukocyte quantity [37]. rs2865303 was a clumped SNP associated with TUD [38] and was previously reported as a significant association of externalizing behavior [39]. Finally, rs865020 was a clumped SNP from MDD [40] with no previously reported associations; it was also not GWS in MDD.

We used FUMA [41] to perform a suite of post-GWAS analyses on PC1. We found that genes significant via a gene-based association test carried out in MAGMA were nominally enriched for brain tissues, but these results were not significant after Bonferroni correction.

Finally, we applied the NNM-sparse model to examine how reproducible the factor structure of our derived **X** matrix was across algorithms. We observed broadly similar clustering behavior and factor structure; further we observed *R*^2^ = 0.611 between **X** derived by NNM-sparse compared to **X** derived by robust PCA (Supplementary Note).

## 3 Methods

### 3.1 Frank-Wolfe algorithms for rank minimization

We use the Frank-Wolfe (FW) algorithm [53–55] to solve the constrained optimization problem. Other methods [56] for solving NNM, including singular value thresholding [27, 57] and proximal methods [58], are computationally more expensive than the FW-type methods. Frank-Wolfe uses significantly less expensive linear subproblems per iteration compared to the quadratic problems in the latter, which can make certain problems intractable; for *e*.*g*., the dual of structural SVMs [59], or an order of magnitude increase in iteration cost for the trace norm (leading eigenvector vs. SVD) [60]. For the FW algorithm, the loss function for the NNM problem is given by,

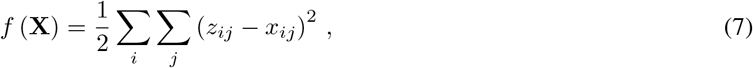

and its gradient is given by,

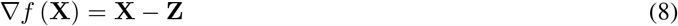

We outline the steps for FW minimization in Algorithm 1. The main bottleneck in implementing the algorithm is solving the linear optimization subproblem in Line 2. The objective of the subproblem is linear even though the constraint set Ω may not be. Since Ω is compact, the solution to the subproblem always exists. After finding a solution **S**_*t*_ to the linear optimization subproblem, the rest of the algorithm concerns finding the appropriate step size to move in the direction dictated by **D**_*t*_: = **X**_*t−*1_ *−* **S**_*t*_. The new iterate is a convex combination of the previous iterate and **S**_*t*_.

The term *g*_*t*_ is called the *surrogate duality gap* which is also a *certificate* of the current approximation quality, *i*.*e*., *g*_*t* 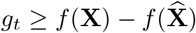_ where 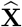 is the desired solution of the optimization. Although 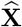 is not known, we can easily compute **X** and *g*_*t*_, which can be used as a convergence criterion.

### Linear optimization subproblem

Denote the singular value decomposition of ▽*f* (**X**_*t−*1_) as **UDV**^T^, where **U** ℝ^*N ×K*^ and **V**∈ ℝ^*P ×K*^ are orthogonal matrices, **U**^T^**U** = **V**^T^**V** = 𝕀_*K*_. The columns of **U** and the columns of **V** are called left-singular vectors and right-singular vectors, respectively. **D** ∈ℝ^*K×K*^ is a diagonal matrix whose diagonal entries are the singular values. The rank of the matrix is *K* = min *{N, P}*. Then the solution to the subproblem in Line 2 is the convex hull of the singular vectors with maximal singular value, appropriately scaled. In practice, we often take the solution 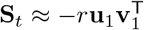, where **u**_1_ and **v**_1_ denote the first columns of **U** and **V** respectively [54, 55].

#### Algorithm 1

Frank-Wolfe algorithm for nuclear norm minimization

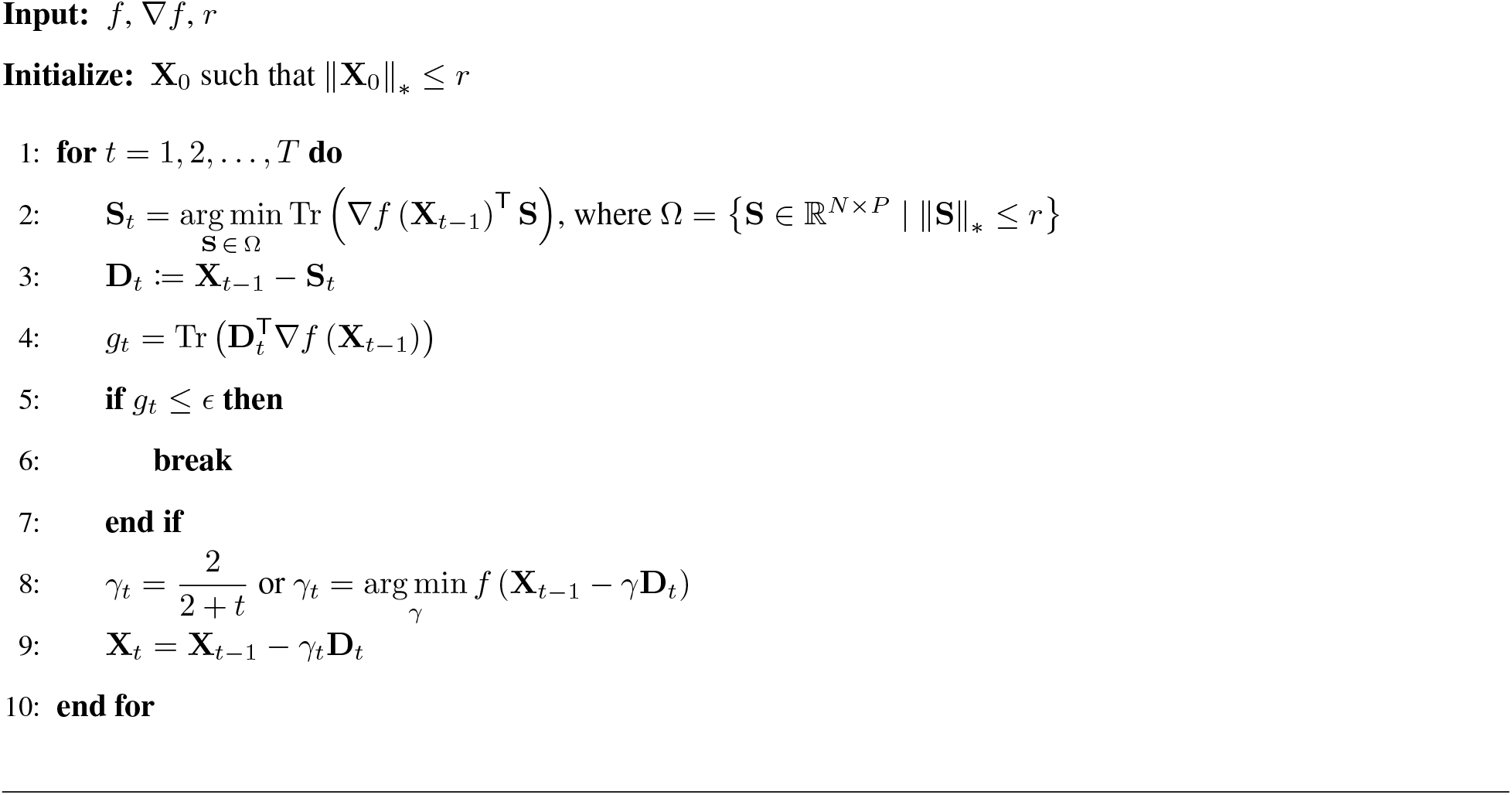

**Step size**. The step size for the subproblem in Line 8 can be derived analytically. Define

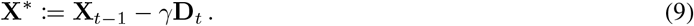

From (7), we have,

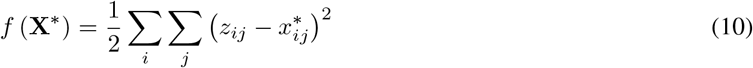

The derivative of this function is,

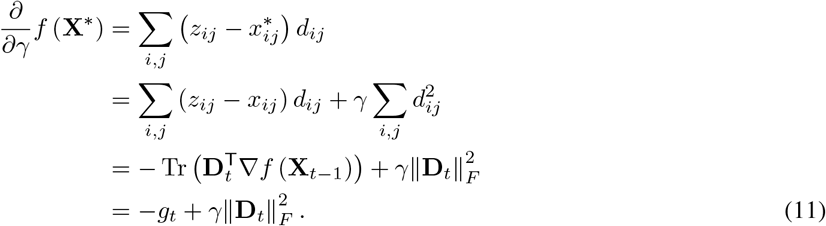

Setting this equal to 0, we obtain the optimum step size as,

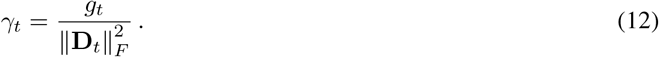

### NNM-Sparse

As discussed in Section 2.1, we want to extract sparse outliers from the input data, and propose our problem as,

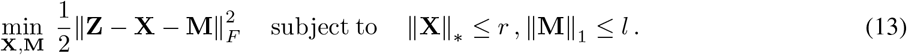

We identify (**X, M**) as the estimand and use the Frank-Wolfe algorithm to solve (13). The loss function and the gradients are given by,

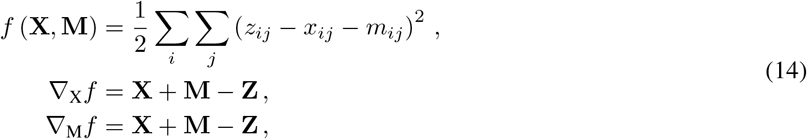

so that the corresponding gradient map can be obtained *∇f* (**X, M**) = (*∇*_X_*f, ∇*_M_*f*). In Algorithm 2, we outline the optimization steps. Here, we need to solve two linear optimization subproblem, corresponding to the two constraints. The nuclear norm regularization for the subproblem in Line 2 is exactly the same as in Algorithm 1. The *𝓁*_1_ norm regularization for the subproblem in Line 3 can be solved by noting that the dual of the *𝓁*_1_ norm is the in *𝓁*_*∞*_ norm, *i*.*e*., ∥**Z**∥_*∞*_: = max_(*i,j*)_ |*z*_*ij*_| = max_∥**X**∥*≤*1_ Tr (**Z**^T^**X**). Therefore, one minimizer is 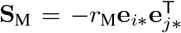 where (*i*, j\**) ∈ arg max_*i,j*_ |*x*_*ij*_ + *m*_*ij*_ *−z*_*ij*_|, *i*.*e*., **S**_M_ has exactly one non-zero element. In the standard FW algorithm, the update for the sparse component updates only one element of the matrix (Line 11). To improve the speed of the algorithm, we include an additional step within each loop to minimize the sparse component, as suggested by Mu *et al*. [61]. Given a threshold *β >* 0, the projection onto the *𝓁*_1_ ball minimizes the sparse component,

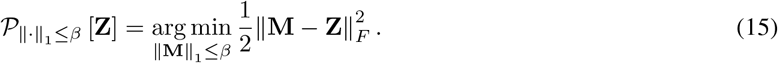

#### Algorithm 2

Frank-Wolfe algorithm for sparse nuclear norm minimization (13)

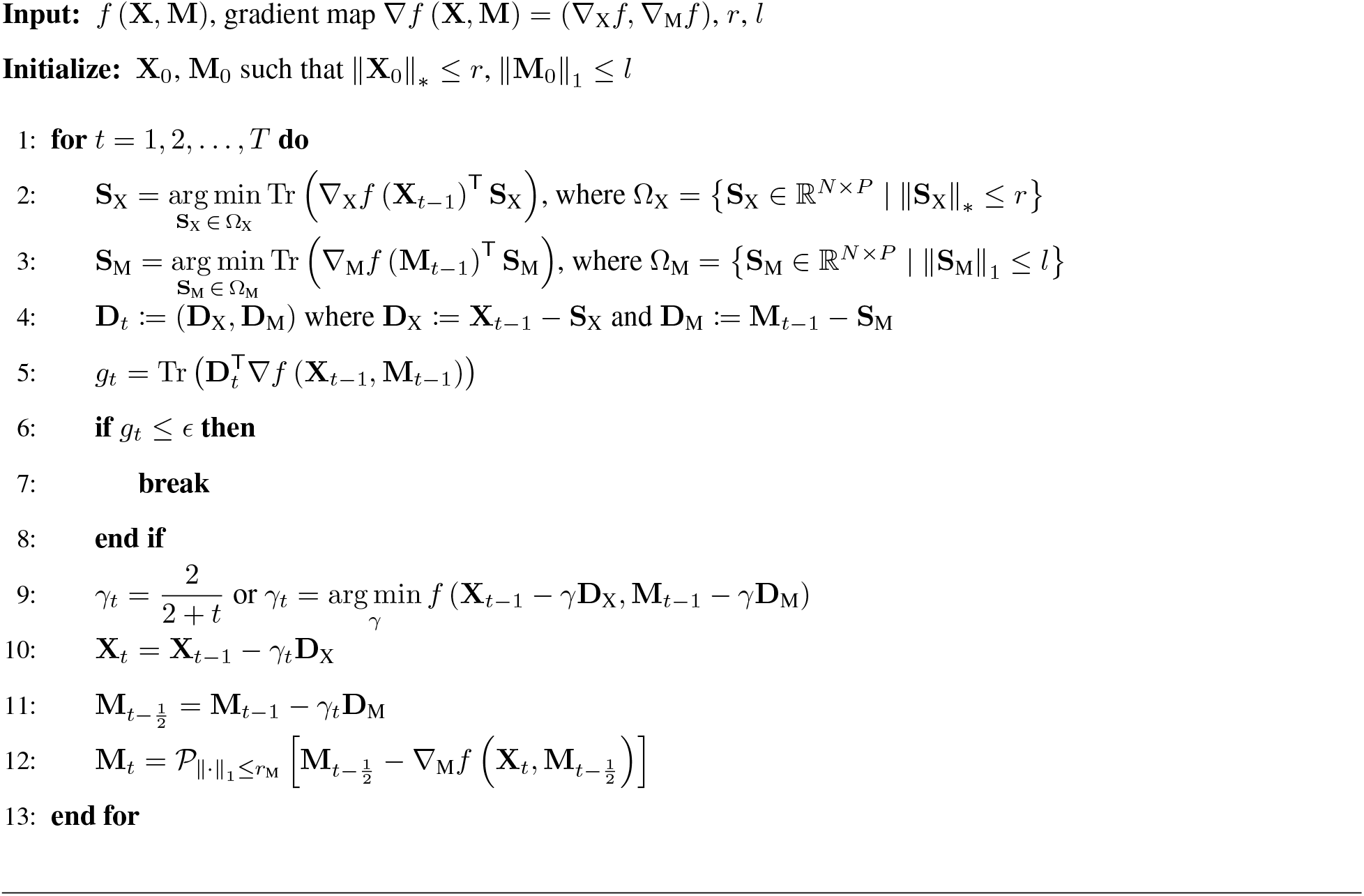

As shown previously [61], the projected gradient step in Line 12 ensures that the objective is less than that produced by the standard FW step, *i*.*e*., 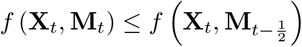.

### 3.2 Application to 14 psychiatric traits

We clumped SNPs per disorder using genotypes from 16,886 European individuals from the HRC as a reference with the following parameters in PLINK1.9: *P <* 5*e*^*−*5^, window = 10, 000kb, *r*^2^ *<* 0.1 [62]. For OUD summary statistics, we obtained the full panel of tested SNPs in European cohorts from the authors [52]. We then annotated SNPs by position and allele using the 1000 Genomes European reference panel (SNP build 151) [63]. We filtered SNPs for missingness across phenotypes, whereby SNPs not present in more than 4 phenotypes were excluded. We verified that SNP values across traits were centered around zero by visualizing their Z-score distributions. We used the Robust PCA algorithm from `Clorinn` with a *λ* value of 0.016 to obtain a solution with approximately 50% non-zero values in *M*. We performed PCA on *X* with 10 PCs using the *scikit-learn* package [64]. Silhoutte scores can be used to index the accuracy of a clustering solution based on comparing the similarity between points within a cluster to those in other clusters. The score can be defined per cluster *k* via

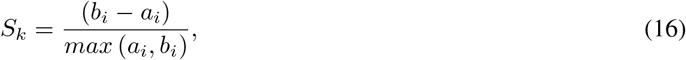

where *a* is the mean intra-cluster distance between sample *i* and members of its assigned cluster, and *b* is the smallest distance between sample *i* and a member of its nearest non-assigned cluster. The output of this score is bounded between *−* 1 and 1, with higher scores indicating less overlapping clusters. Distances between points were measured in the euclidean space. Results reported are the mean Silhouette score across samples per solution.

We obtained cos^2^ scores via

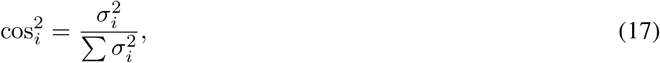

where *σ* is the standard deviation of PC *i*.

We tested for a difference in means between numbers of traits where SNPs are significant across sets using an independent t-test of means. We calculated pleiotropy scores using a Python implementation of [34] in ‘magnitude’ mode, and tested for a difference in means between SNP sets using an independent t-test of means. We did not correct the score for LD as minimal differences were reported in the original paper when carrying out this operation.

We derived an association test statistic per SNP for their association with PC values using the procedure described in [35]. Briefly, we performed a singular value decomposition of the input *Z* matrix to obtain the right singular vectors *V* ^*T*^. The PC score is the product of the input data multiplied by the right singular vectors, yielding a *N × N* matrix (where *N* is the number of traits). The association score was obtained by regressing the scaled values of PC 1 from this matrix across phenotypes on the SNP association Z-scores across phenotypes. We repeated this operation for every SNP and used *FUMA* [41] to carry out functional enrichment of GWAS results.

## 4 Discussion

`Clorinn` showed comparable or better performance to similar methods for factor discovery in a range of simulation scenarios (Figure 2). This is likely due to the fact that `Clorinn` first applies LRMA before factor discovery to derive an outlier-free version of the input matrix. This added step ensures that subsequent analyses are not confounded by large amounts of noise, or the addition of extra phenotypes/variants. This property is likely to be useful for consortium-level analyses, whereby re-analysis with additional cohorts is required. Further, the Frank-Wolfe convex optimization procedure guarantees reproducibility of the **X** matrix regardless of initialization. This convexity feature is attractive from a solution stability and reproducibility standpoint. Secondly, we find that hidden factors derived via `Clorinn` can be used to accurately cluster large groups of diverse phenotypes in a biobank-scale application, as evidence from our tSNE embedding plot of 30 broad phenotype categories (Figure 3). Of particular note is the large groupings of dietary intake and mental health traits respectively (light blue and brown points in Figure 3). This suggests that granular categorical distinctions of phenotypes are possible even when considering large numbers of traits. In a phenotype-focused example, we found PC26 as the factor loading on Type 2 diabetes to the largest degree. An examination of the underlying biology via squared cosine scores establishes good concordance with previous literature on tissue enrichment and associated genes (Figure 4). For example, the top variant for PC26 was mapped to *TCF7L2*, which has been previously identified as a significant GWAS association of the condition [65]. We also observe that 4 of the top 8 variants for PC26 map to *CETP*, a plasma protein responsible for cholesterol transport. Recent research has described the potential of *CETP* inhibitors for the treatment of new-onset diabetes, demonstrating that our analysis can identify variants with therapeutic potential [66]. We can also leverage the fact that traits can be loaded on by multiple factors for biological insight, as evidenced through Figure 4B, whereby we examined variable traits along PC18 and PC26, which are the 2 top factors of Type 2 Diabetes. We find that several blood traits have large variance in this plot, including albumin/globulin ratio and white blood cell leukocyte count. Unsurprisingly, these traits have been previously reported as having significant genetic correlations with diabetes [67]. Additionally, we find that S-LDSC results implicate the pancreas as the main tissue of interest, which is relevant given that the molecular basis of Type 2 diabetes originates in this tissue [68] (Figure 4D). These experiments verify that biologically meaningful factors can be identified from biobank-scale data in a computationally tractable and reproducible manner. Furthermore, it demonstrates that biological discovery is possible at a broad scale for the identification of large category groupings, and at a more granular level via a focus on specific phenotype-factor relationships.

Additionally, we find that `Clorinn` recapitulates gSEM results in our psychiatric trait application. Firstly, phenotypes are shown to group based on their first two PCs according to categories defined in previous work after running PCA on the resultant **X** matrix [33] (Figure 5A). This is interesting given that `Clorinn` makes use of information from just 5, 815 SNPs per trait. Furthermore, we found that the mean Silhouette score was improved via application of `Clorinn`, suggesting that the accuracy of cluster resolution is increased through derivation of an “outlier-free” SNP matrix. Our squared cosine embeddings describing loading of a factor on traits showed good agreement with results of the 5-factor model from [33] (Figure 5B). Specifically, we found that all factors from the 5-factor gSEM model are represented in the cos^2^ score embeddings in at least one PC. An advantage of this approach is the reduced amount of SNP information required per phenotype; additionally, a hypothesized phenotype factor structure is not required. This is useful from a computational and factor discovery standpoint. For instance, we find that internalizing disorders and compulsive disorders load primarily on PC1 in Figure 5B. We also observe that categories tend to contribute to PC activity primarily in pairs. For example, internalizing disorders in PC1 and PC6, and SCZ/BD and substance use disorders in PC2 and PC8. Further, certain phenotype categories have more specific contribution patterns to factors, such as in neurodevelopmental disorders, which contributes largely to PC4 and PC10, but with no discernible category pairing. This visualization technique has great potential for identifying correlated sets of traits belonging to broader groupings, especially where the number of traits is large. An advantage of `Clorinn` from this standpoint is its potential in both biobank-scale factor analysis and more controlled applications. Comparable methods are limited to the derivation of factors from larger numbers of phenotypes. We have demonstrated that `Clorinn` can be applied to thousands of of traits.

We also demonstrate the discovery potential of `Clorinn` in our psychiatric application via statistical hypothesis testing of factor loadings and the definition of pleiotropic and disorder-specific SNP sets. We found that 18 SNPs were GWS after correlating variant *Z* scores to factor activity across phenotypes, with 4 variants reported which were not GWS in any of the top 4 phenotypes loaded on by PC1. This framework has the potential to obtain proxy genetic associations with unobserved latent variables, which is a conceptually similar goal to approaches such as gSEM. Our approach is an attractive alternative owing to the computational ease of analysis and the sparsity of genetic information required. The definition of disorder-specific SNP sets could also be used to describe properties related to the residual variance per phenotype from gSEM and FactorGO, although an application of this was not explored in this work.

## 5 Limitations

Our simulation results did not include a direct comparison of our method to *GLEANR*. Our work also did not seek to define residual variance of phenotypes, but the information returned by `Clorinn` could be used for this application. Finally, while our LRMA approach has many useful properties, our resultant factors do not have a direct SNP-mapping (except through the outlier-free 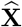). This abstraction may makes it more difficult to interpret associated factors and their constituent elements. Similarly, an important point of future research will be developing a thorough interpretation of SNP 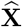 values relative to the input SNP-trait associations.

It has recently been shown that assortative mating inflates estimates of genetic correlation[69]. Factorization approaches like `Clorinn` will be influenced by this inflation, and it remains an open question how such analyses might be appropriately corrected.

## 6 Conclusion

Here, we have developed a computationally efficient and flexible modeling approach to describe robust latent genetic factors of an arbitrary number of GWAS results. Our method is reproducible and does not constitute a direct analysis of the traits-effects matrix for the definition of latent factors, which has several attractive properties. We anticipate that `Clorinn` can be used for analysis at the trait and variant level in a variety of applications, especially as the number of phenotypes scale.

## Data Availability

All data is publicly available from the cited studies.

## 7 Code availability

All code is publicly available at https://github.com/banskt/colormann.

## 8 Acknowledgments

Research reported in this manuscript was supported by the National Institute of Mental Health of the National Institutes of Health under award number R01MH130879. The content is solely the responsibility of the authors and does not necessarily represent the official views of the National Institutes of Health.

## Supplementary note

**Figure S1:**
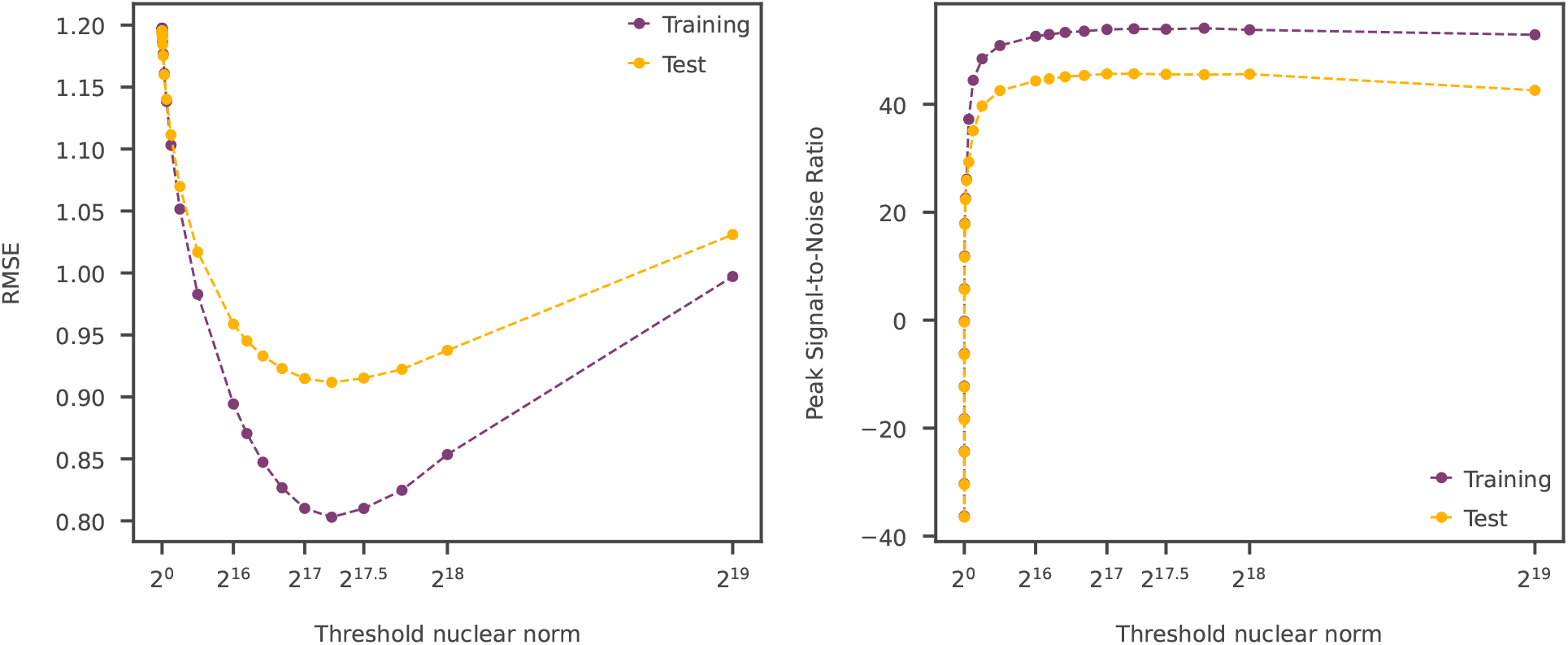
Cross-validation to obtain rank threshold for NNM-Sparse FW algorithm on PanUKB data. Both error metrics, root mean square error (RMSE, left panel) and peak signal-to-noise ratio (right panel) suggests a threshold of *r* = 2^17.25^ = 155872

**Figure S2:**
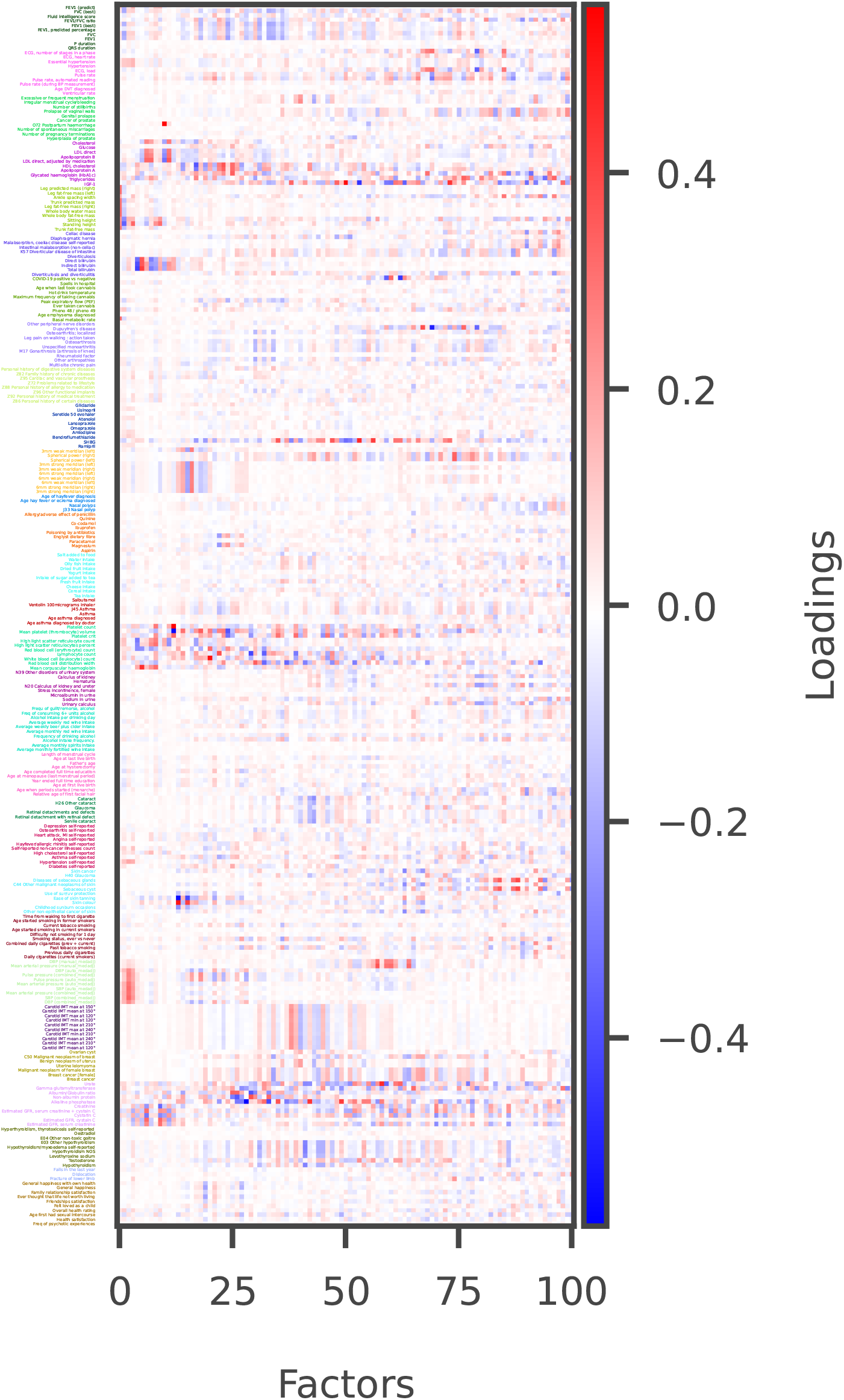
Contribution of some selected traits (shown on y-axis) on the top 100 factors (shown on x-axis) obtained from the application of Clorinn on the PanUKB dataset. The heatmap shows the magnitude of the loadings (color scale shown on the right). The traits were selected based on their estimated heritability (*h*^2^ *≥* 0.001), and a maximum of 10 traits were retained for each text-based disease category, see Figure 3 in main text.

**Figure S3:**
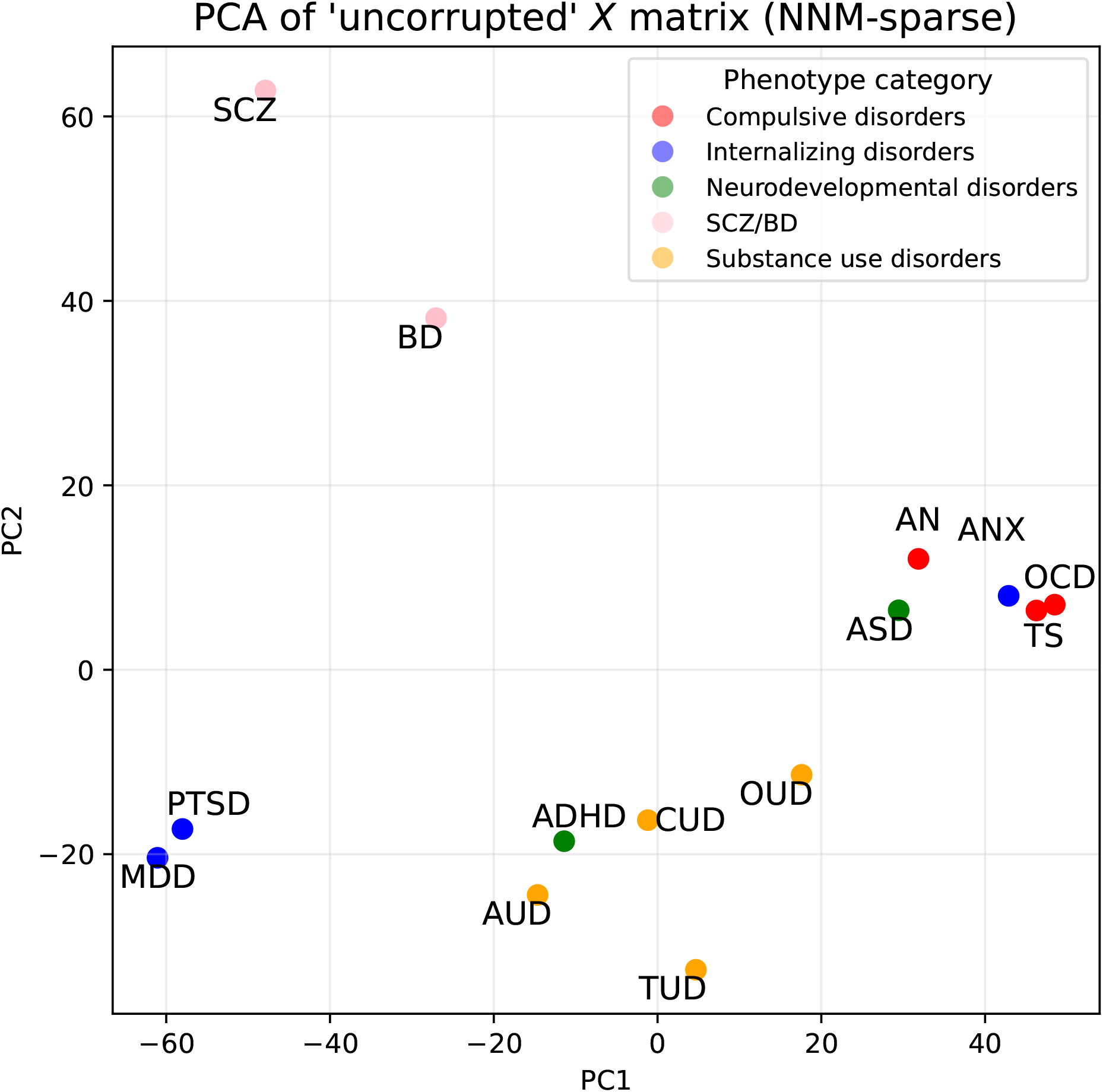
PC1 and PC2 of NNM-sparse-derived *X* for the application of Clorinn to 14 psychiatric traits. The nuclear norm constraint was chosed with a 5 fold cross validation procedure. Points are colored by their phenotype categories as described previously.

**Figure S4:**
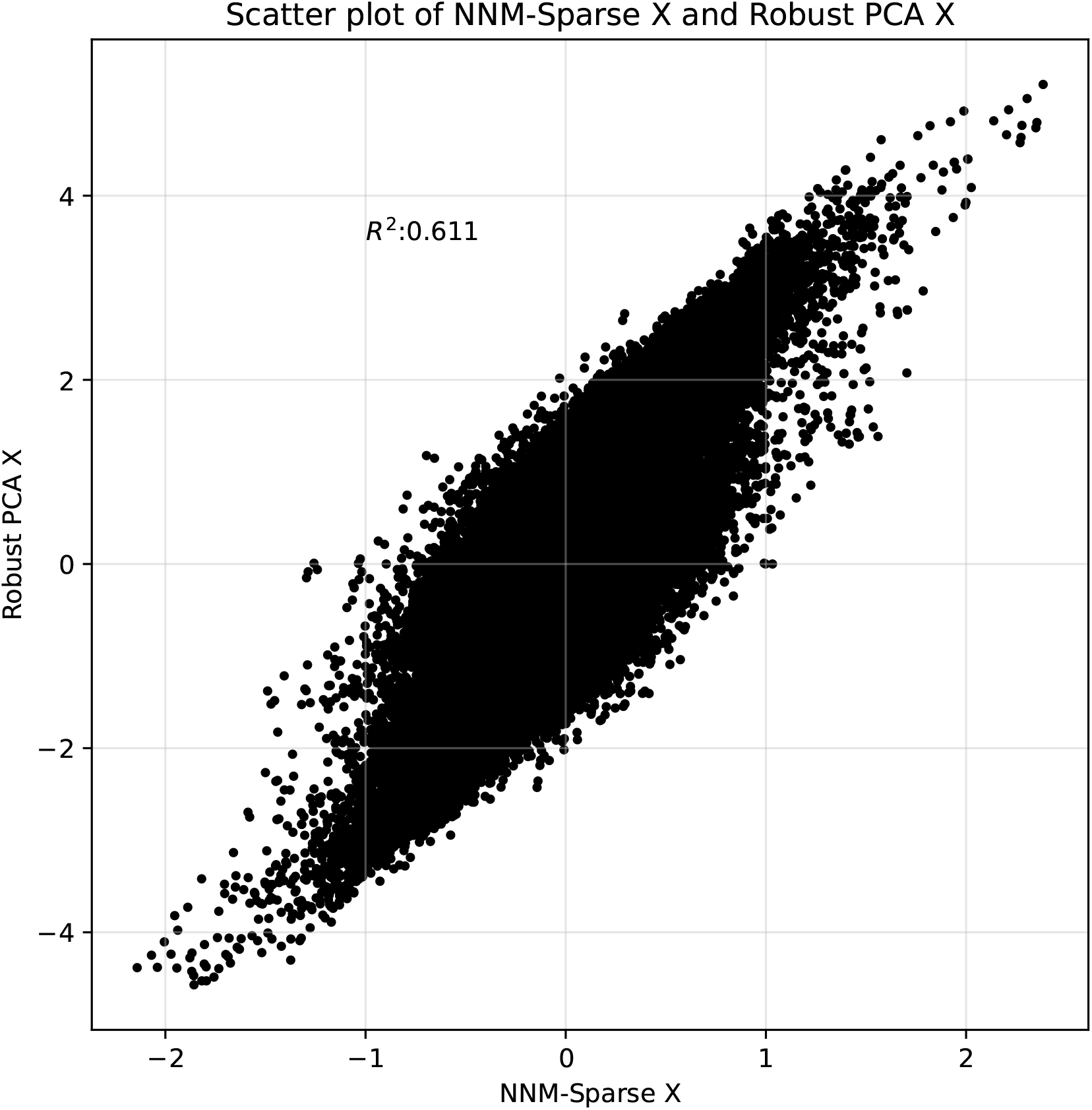
Scatter plot of *X* derived from robust PCA vs. *X* derived using NNM-sparse.

**Figure S5:**
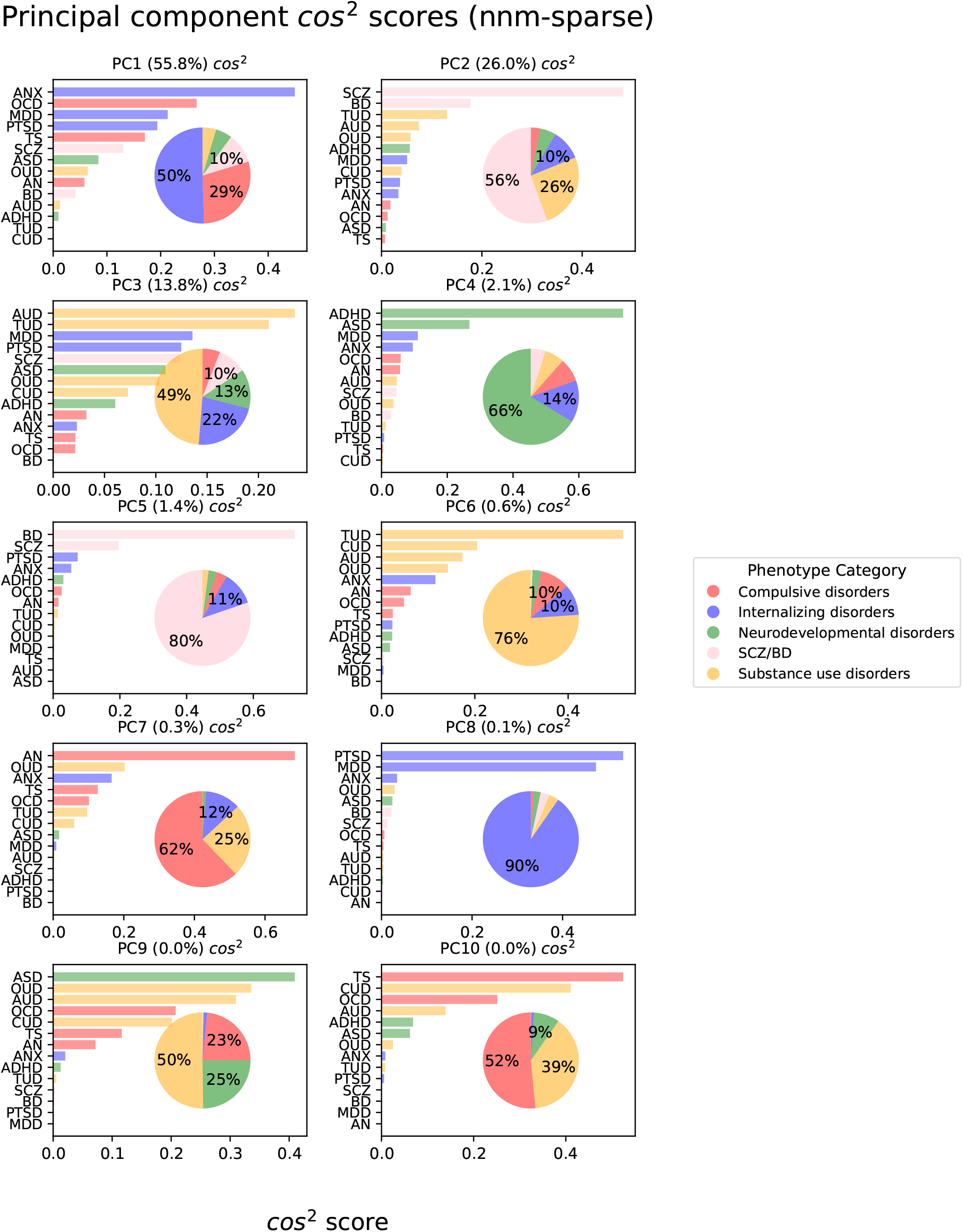
Squared cosine scores for factors derived from NNM-sparse *X*. Legend and details are inherited from Figure 5.

**Figure S6:**
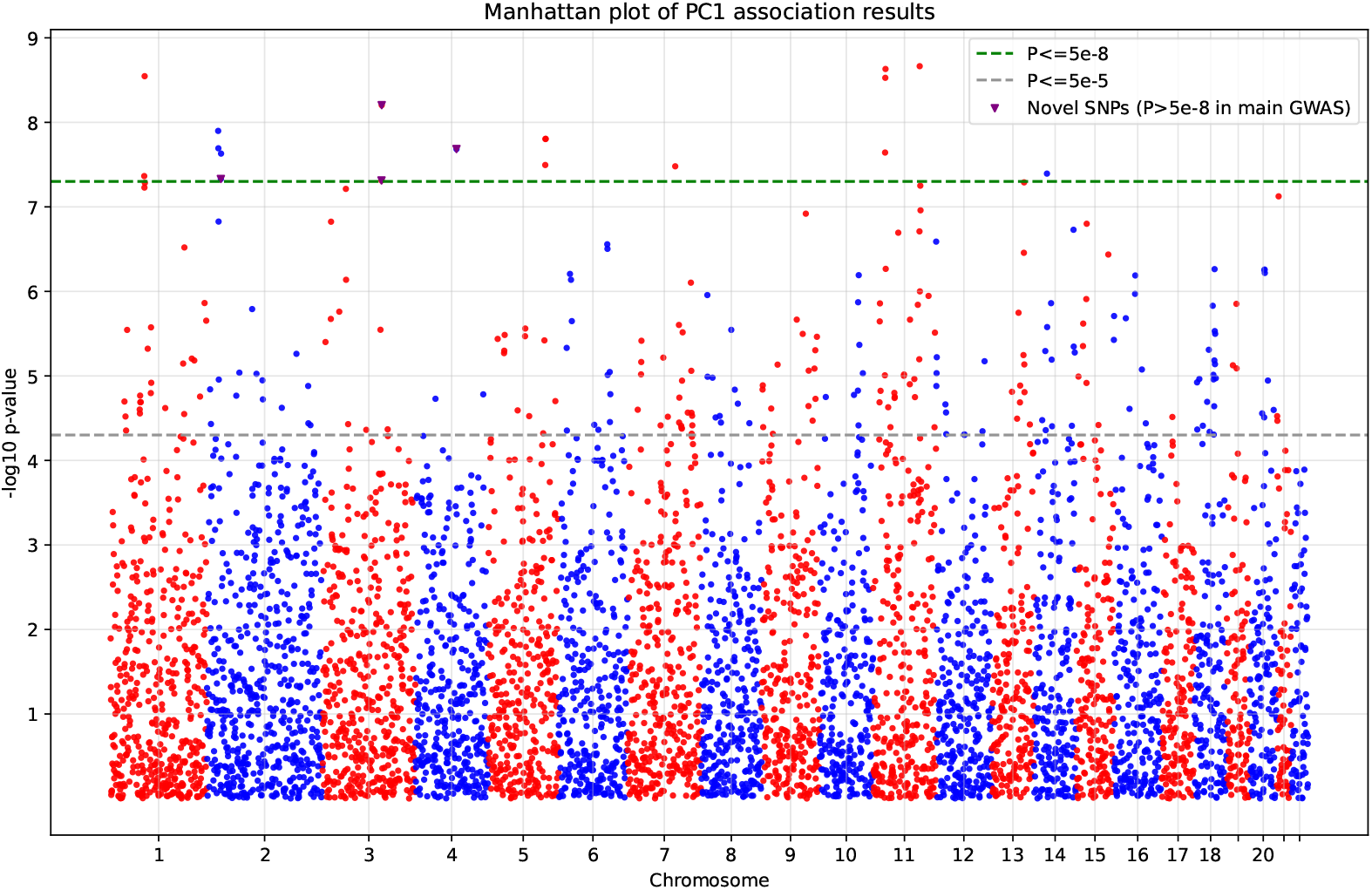
Manhattan plot of PC1 association statistics for 5, 815 SNPs from the application of Clorinn with the robust PCA algorithm. Statistics were derived by regressing PC score per phenotype against raw variant values across phenotypes from the input. Nominal and genome-wide significance lines are drawn with dashes; SNPs found to be GWS for PC1 activity but were not GWS for any of the top 4 loaded phenotypes (MDD, PTSD, OCD, and TS) for PC1 are colored in purple and marked with a triangle. These SNPs are discussed in the main text.

